# Evaluation of a Machine Learning-guided Strategy for Elevated Lipoprotein(a) Screening in Health Systems

**DOI:** 10.1101/2024.03.02.24303582

**Authors:** Arya Aminorroaya, Lovedeep S Dhingra, Evangelos K Oikonomou, Rohan Khera

**Affiliations:** Section of Cardiovascular Medicine, Department of Internal Medicine, Yale School of Medicine, New Haven, CT, USA; Center for Outcomes Research and Evaluation (CORE), Yale New Haven Hospital, New Haven, CT, USA; Section of Health Informatics, Department of Biostatistics, Yale School of Public Health, New Haven, CT, USA

**Author notes:** Corresponding Author: Rohan Khera, MD, MS, 195 Church St, 6^th^ Floor, New Haven, CT 06510;**; @rohan_khera**.

**Keywords:** Lipoprotein(a), Screening, Machine Learning, Electronic Health Records, Preventive Medicine

## Abstract

**Background:** While universal screening for Lp(a) is increasingly recommended, fewer than 0.5% of the patients undergo Lp(a) testing. Here, we assessed the feasibility of deploying Algorithmic Risk Inspection for Screening Elevated Lp(a) (ARISE), a validated machine learning tool, to health system EHRs to increase the yield of Lp(a) testing.

**Methods:** We randomly sampled 100,000 patients from the Yale-New Haven Health System (YNHHS) to evaluate the feasibility of ARISE deployment. We also evaluated Lp(a) tested populations in the YNHHS (N=7,981) and the Vanderbilt University Medical Center (VUMC) (N=10,635) to assess the association of ARISE score with elevated Lp(a). To compare the representativeness of the Lp(a) tested population, we included 456,815 participants from the UK Biobank and 23,280 from three US-based cohorts of ARIC, CARDIA, and MESA.

**Results:** Among 100,000 randomly selected YNHHS patients, 413 (0.4%) had undergone Lp(a) measurement. ARISE score could be computed for 31,586 patients based on existing data, identifying 2,376 (7.5%) patients with a high probability of elevated Lp(a). A positive ARISE score was associated with significantly higher odds of elevated Lp(a) in the YNHHS (OR 1.87, 95% CI, 1.65-2.12) and the VUMC (OR 1.41, 95% CI, 1.24-1.60). The Lp(a) tested population significantly differed from other study cohorts in terms of ARISE features.

**Conclusions:** We demonstrate the feasibility of deployment of ARISE in US health systems to define the risk of elevated Lp(a), enabling a high-yield testing strategy. We also confirm the very low adoption of Lp(a) testing, which is also being restricted to a highly selected population.

## INTRODUCTION

Lipoprotein(a) [Lp(a)], a novel biomarker of atherosclerotic cardiovascular disease (ASCVD), is increasingly being recommended to be measured once in a lifetime for everyone.^1–6^ Elevated Lp(a) is associated with an 11% higher hazard of ASCVD development per 50 nmol/L increment and should be treated with intensive cardiovascular risk reduction strategies, including lipid-lowering medications and lifestyle modification.^5^ Additionally, the advent of novel targeted therapies leading to large reductions in serum levels of Lp(a) can transform the treatment of individuals with elevated Lp(a), especially if ongoing phase III randomized clinical trials demonstrate a reduction of adverse cardiovascular outcomes by these agents.^7–14^ Despite these developments, fewer than 0.5% of the patients undergo Lp(a) testing.^15–17^

To address the limited testing of Lp(a) in contemporary practice, we have developed and validated a novel machine learning model to increase the yield of Lp(a) testing using six features, including ASCVD history, use of statins, use of anti-hypertensive medications, serum low-density lipoprotein cholesterol (LDL-C), serum high-density lipoprotein cholesterol (HDL-C), and serum triglycerides.^18^ The model, Algorithmic Risk Inspection for Screening Elevated Lp(a) (ARISE), demonstrated a consistent reduction of the number-needed-to-test to find one case with elevated Lp(a) by more than 50% in four multinational cohorts. Given the computability of ARISE’s features in electronic health records (EHR), deployment of ARISE in health systems with subsequent testing of screen-positive patients has the potential to identify a large number of patients with elevated Lp(a) at scale.^18^ However, the feasibility of its deployment in EHR and its implications for Lp(a) screening have not been assessed previously.

In this study, we sought to assess the feasibility of ARISE deployment in two large and diverse US health systems. In these health systems, we also compared the representation of the population whose Lp(a) levels were tested against large population-based cohorts in the US and the UK, and against EHR patients in one of the health systems who had not undergone Lp(a) testing.

## METHODS

Detailed methods are available in the **Supplemental Material**. The Yale Institutional Review Board approved the study protocol and waived the need for informed consent as the study involves analyzing pre-existing data. Patients who opted out of research studies at the Yale-New Haven Health System (YNHHS) were not included in the study. The use of Vanderbilt University Medical Center (VUMC) data was classified as non-human subject research by VUMC’s Institutional Review Board as the study involves analyzing de-identified records. This research has been conducted using the UK Biobank (UKB) Resource under Application Number #71033. The participants of Atherosclerosis Risk in Communities (ARIC), Coronary Artery Risk Development in Young Adults (CARDIA), and Multi-Ethnic Study of Atherosclerosis (MESA) provided informed consent for participation, and their de-identified data were analyzed in this study. A secure workspace on the American Heart Association Precision Medicine Platform was used for data analysis.

## RESULTS

### Study Population

In the randomly selected 100,000 patients drawn from the YNHHS who had at least one healthcare encounter, the median age of individuals was 50 [IQR, 31-65] years and 55,347 (55.4%) were women (**Table 1**). In the separate cohort of 7,981 individuals who had undergone Lp(a) assessment in YNHHS, the median age was 57 [46-66] years, and 4,534 (56.8%) were women compared with a corresponding cohort of 10,635 patients in the VUMC with Lp(a) testing with a median age of 46 [17-59] and 5,037 (47.4%) women. In the UKB, 456,815 participants received Lp(a) testing with a median age of 58 [51-64] years and 247,511 (54.2%) women compared with 23,280 participants in the US cohorts with a median age of 53 [46-60] years and 12,572 (54.0%) women.

**Table 1.** Baseline Characteristics of the Study Population.

| Characteristic | YNHHS Sample | YNHHS Sample LDL-C/Lp(a) Tested | YNHHS Lp(a) Tested | VUMC Lp(a) Tested | UK Biobank | US Cohorts |
| --- | --- | --- | --- | --- | --- | --- |
| **Number** | 100,000 | 34,927 | 7,981 | 10,635 | 456,815 | 23,280 |
| **Age (years)** | 50 [31-65] | 56 [40-68] | 57 [46-66] | 46 [17-59] | 58 [51-64] | 53 [46-60] |
| **Female Sex** | 55347 (55.4%) | 18936 (54.2%) | 4534 (56.8) | 5037 (47.4%) | 247511 (54.2%) | 12572 (54.0) |
| **Race and Ethnicity\*** |  |  |  |  |  |  |
| Hispanic | 11468 (12.9%) | 4102 (12.1%) | 606 (7.9%) | 511 (5.4%) | - | 1062 (4.6%) |
| White | 61659 (69.4%) | 23906 (70.5%) | 6002 (77.8%) | 7867 (82.9%) | 430601 (94.7%) | 14707 (63.2%) |
| Black | 9288 (10.5%) | 4144 (12.2%) | 640 (8.3%) | 820 (8.6%) | 7079 (1.6%) | 6952 (29.9%) |
| Asian | 2871 (3.2%) | 978 (2.9%) | 240 (3.1%) | 192 (2.0%) | 1418 (0.3%) | 559 (2.4%) |
| Other | 3502 (3.9%) | 785 (2.3%) | 226 (2.9%) | 97 (1.0%) | 15581 (3.4%) | 0 (0%) |
| **ASCVD** | 6928 (6.9%) | 5444 (15.6%) | 3354 (42.0%) | 3404 (32.0%) | 22534 (4.9%) | 2649 (11.4%) |
| **Statin** | 7497 (7.5%) | 6442 (18.4%) | 2950 (37.0%) | 4285 (40.3%) | 76292 (16.7%) | 95 (0.4%) |
| **Antihypertensive Medication** | 6594 (6.6%) | 5537 (15.9%) | 3249 (40.7%) | 4348 (40.9%) | 96182 (21.2%) | 6034 (26.0%) |
| **LDL-C (mg/dL)** | - | 99.0 [77.4-124.0] | 105.0 [78.0-134.0] | 124.0 [90.0-161.0] | 135.9 [113.7-159.2] | 126.0 [102.5-152.2] |
| **HDL-C (mg/dL)** | - | 53.0 [43.0-66.0] | 56.0 [44.0-69.0] | 45.0 [37.0-55.0] | 54.1 [45.3-64.8] | 49.1 [40.0-60.7] |
| **Cholesterol (mg/dL)** | - | 178.0 [152.0-207.0] | 187.0 [154.0-220.0] | 209.0 [171.0-253.0] | 218.5 [189.6-248.5] | 201.0 [176.0-230.0] |
| **Triglycerides (mg/dL)** | - | 100.0 [71.0-145.0] | 98.0 [70.0-145.0] | 142.0 [91.0-233.0] | 131.4 [92.8-190.3] | 101.0 [70.0-148.0] |

**Abbreviations:** ASCVD, atherosclerotic cardiovascular disease; HDL-C, high-density lipoprotein cholesterol; LDL-C, low-density lipoprotein cholesterol; Lp(a), Lipoprotein(a); YNHHS, Yale-New Haven Health System.
\* Race and ethnicity are reported as a combination, where 'White' denotes non-Hispanic White individuals, 'Black' refers to non-Hispanic Black individuals, 'Asian' signifies non-Hispanic Asian individuals, and 'Other' describes non-Hispanic individuals identifying with a race other than White, Black, or Asian.

In the random 100,000 sample from the YNHHS, 10.5% identified as non-Hispanic Black and 12.9% as Hispanic. Among the Lp(a) tested population, YNHHS had 8.3% non-Hispanic Black and 7.9% Hispanic individuals compared with 8.6% and 5.4%, respectively, in the VUMC. This contrasts with the population-based cohorts where 1.6% in the UKB and 29.9% in the US cohorts identified as non-Hispanic Black. While Hispanic ethnicity was not represented in the UKB, the US cohorts included 4.6% Hispanic participants. A history of ASCVD, and statin and antihypertensive medication use were more prevalent in the YNHHS and VUMC patients with Lp(a) assessment compared with the YNHHS random sample, UKB, and the US-based cohorts (**Table 1**).

### Feasibility of ARISE Deployment in the EHR

Among the 100,000 randomly selected YNHHS patients, 413 (0.4%) patients had undergone at least one Lp(a) measurement of whom 85 (20.6%) had an elevated Lp(a) (**Figure 1**). Of the remaining patients, 34,514 (34.7%) had undergone at least one LDL-C measurement. In this subset, 31,586 (91.5%) patients had data points for all ARISE features. However, 66 (0.2%) patients had missing serum HDL-C, 44 (0.1%) had missing triglycerides, and 2,818 (8.2%) had missing HDL-C and triglycerides within 30 days before their LDL-C measurement in the EHR. In the subset of 31,586 patients with complete ARISE features, 2,376 had a positive ARISE score, indicating a high probability of elevated Lp(a). Thus, 7.5% of the patients with complete data, representing 2.4% of the initial random patient sample, show a high probability of elevated Lp(a). After leveraging ARISE to impute missing data for the remaining patients, we identified 2,451 patients with a positive ARISE score. This represents 7.1% of the patients assessed for LDL-C and 2.5% of the initial random sample.

**Figure 1.**
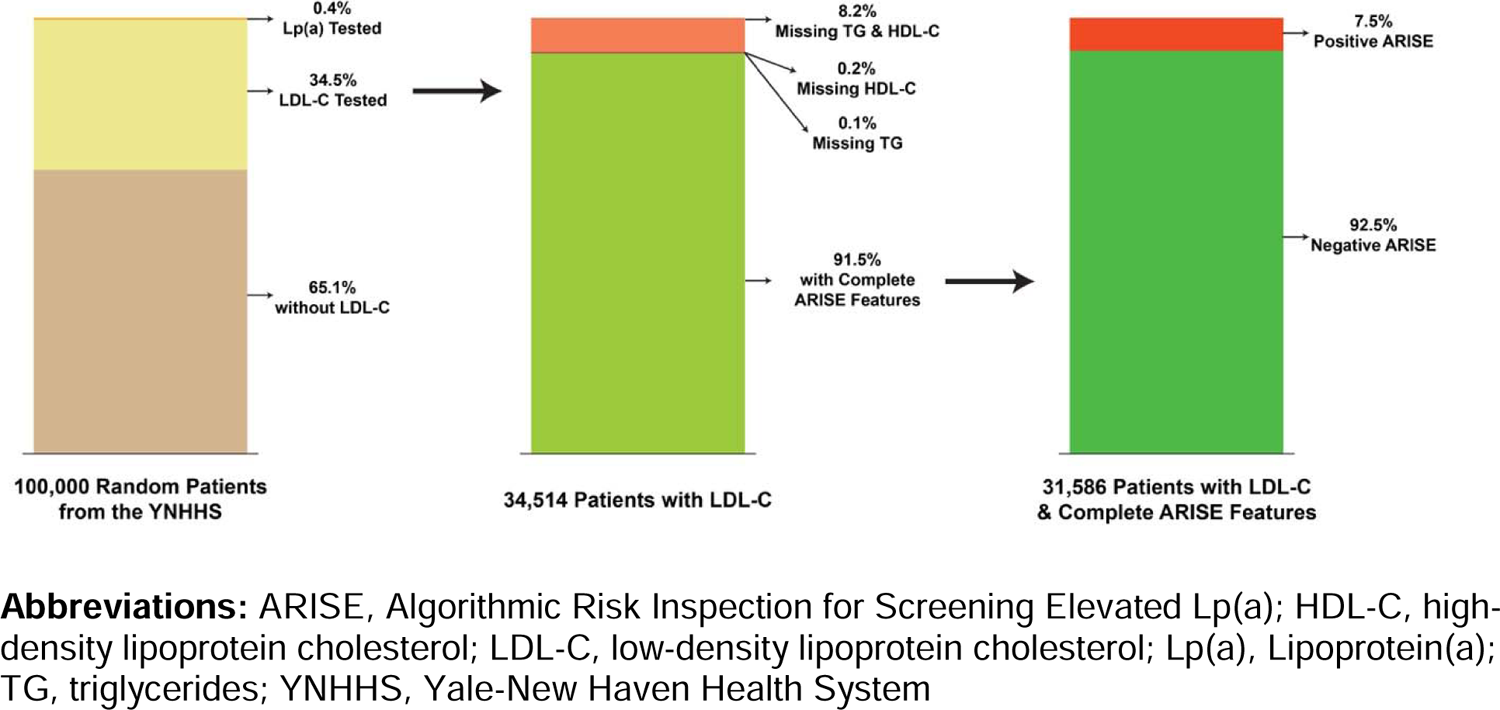
ARISE Deployment in a Random Sample of 100,000 Patients from the Yale-New Haven Health System

### ARISE Deployment in the Lp(a) Tested EHR Populations

Of 7,981 YNHHS patients with Lp(a) testing, 6,330 (79.3%) had a high probability of elevated Lp(a) and 1,480 (18.5%) had elevated Lp(a). In comparison, among 10,635 patients with Lp(a) testing in the VUMC, 3,019 (28.4%) had a high ARISE score and 1,225 (11.5%) had elevated Lp(a). A positive ARISE score was associated with significantly higher odds of elevated Lp(a) in the YNHHS (OR 1.87, 95% CI, 1.65 to 2.12) and the VUMC (OR 1.41, 95% CI, 1.24 to 1.60). Odds of elevated Lp(a) increased by 38% (OR 1.38, 95% CI, 1.31 to 1.46) in the YNHHS and 24% (OR 1.24, 95% CI, 1.17 to 1.31) in the VUMC per one SD increment in the ARISE score.

### Representation of the Lp(a)-tested EHR Populations Compared with Other Study Cohorts

The average Gower’s distances between each pairwise comparison of random YNHHS sample, UKB, and US cohorts ranged from 0.048 [0.039-0.19] to 0.070 [0.055-0.21], representing median and IQR values drawn from these comparisons (**Figure 2**). Nevertheless, it ranged from 0.21 [0.20-0.22] to 0.22 [0.06-0.38] for comparison of the YNHHS cohort with Lp(a) testing against random YNHHS sample, UKB, and US cohorts. Uniform manifold approximation and projection (UMAP) representation of the pairwise comparisons of the study cohorts demonstrated overlap between random YNHHS sample, UKB, and US cohorts while showing separation between the YNHHS cohort with Lp(a) testing and other study cohorts (**Figure 3**).

**Figure 2.**
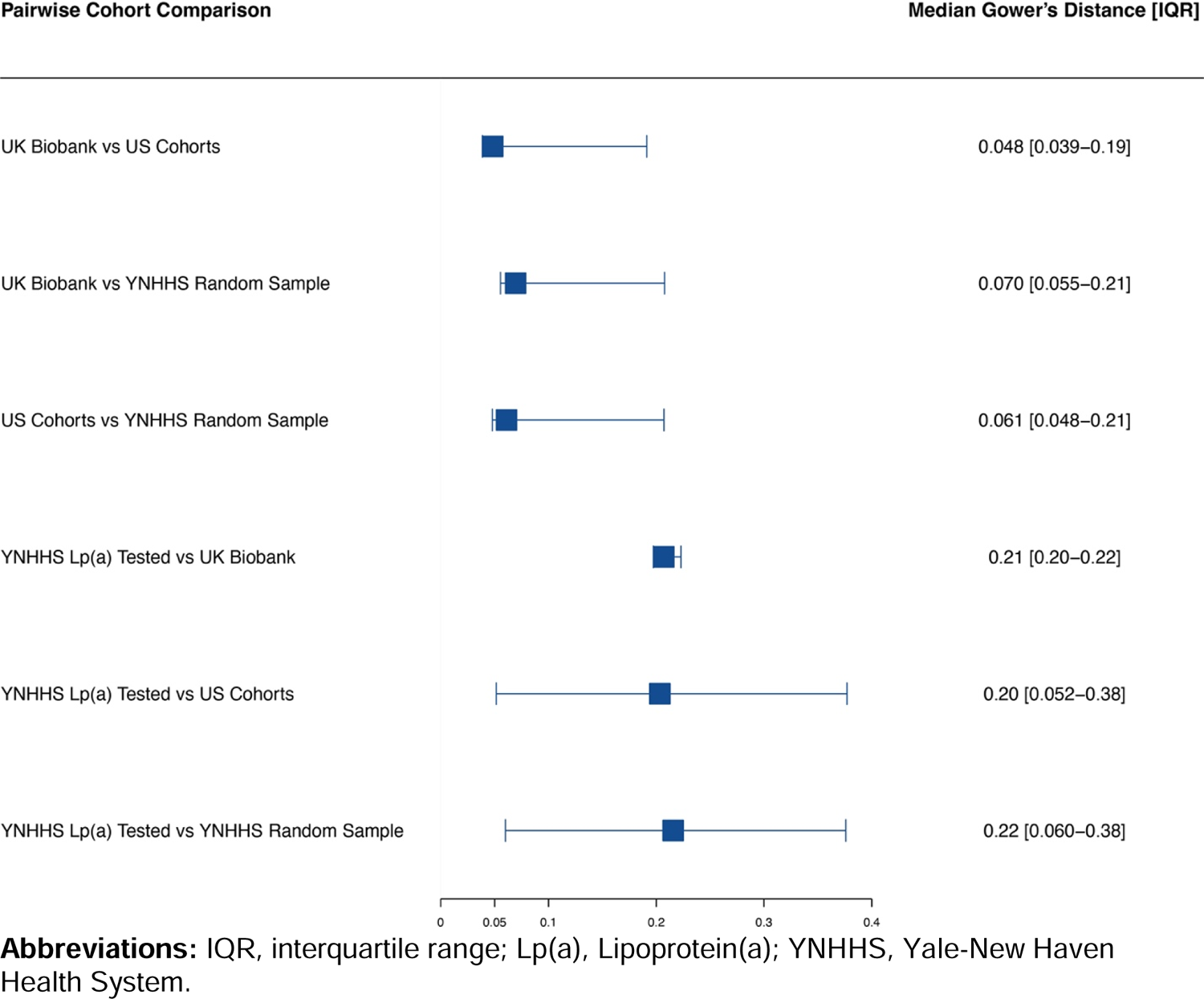
The Median of the Medians of Gower’s Distances Between Individuals Across Each Pairwise Cohort Comparison

**Figure 3.**
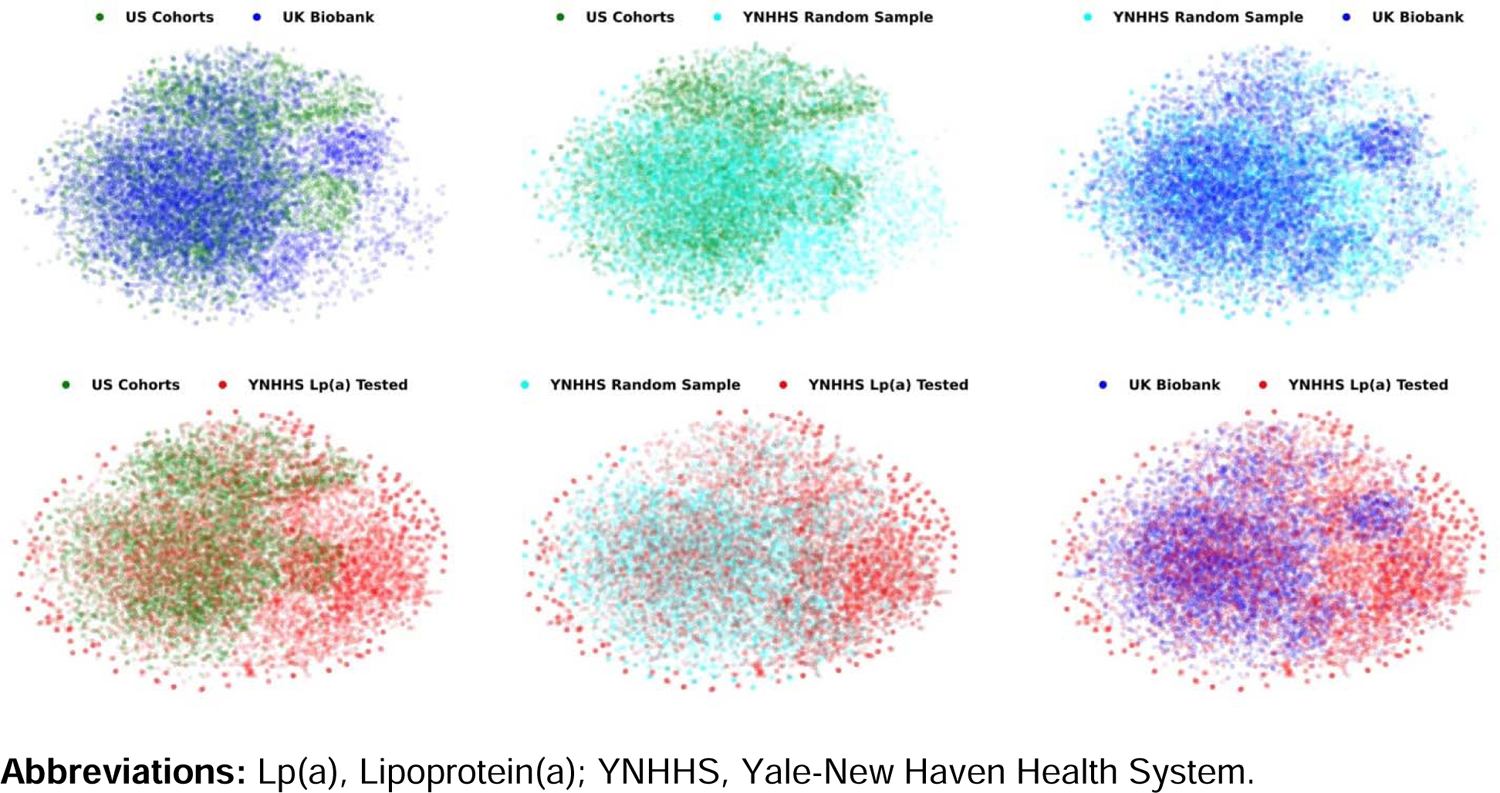
UMAP Representation of Gower’s Distances Between Across Each Pairwise Cohort Comparison

## DISCUSSION

In two large and diverse US health systems, we found that the ARISE score is computable for one-third of the EHR population, demonstrating the feasibility of its deployment as a strategy to increase the yield of Lp(a) testing and enhance identification of those with elevated Lp(a) at scale in the health systems. This is in contrast to 0.4% of the population who underwent Lp(a) testing. A screen-positive ARISE was associated with a 1.5- to 2-fold increased odds of having elevated Lp(a). Our findings underscore the markedly low adoption of Lp(a) testing and demonstrate that the cohort undergoing Lp(a) assessment in the contemporary practice constitutes a highly selective group, in contrast to prevailing guidelines advocating for comprehensive screening across the entire population.

This study builds upon the current literature by demonstrating the feasibility of deployment of a novel algorithmic strategy in a large and diverse US health system for increasing the yield of Lp(a) testing.^18^ Despite the clinical significance of elevated Lp(a) and current guideline recommendations for once-in-a-lifetime universal screening for this condition, only 0.4% have undergone Lp(a) testing which aligns with the reported measures from other health systems and claims databases.^15–17^ Adherence to guideline recommendations in real-world clinical practice remains a challenge in different domains of medicine and cardiology with implementation science being an active area of interest and research.^19,20^ The feasibility of deploying ARISE within health systems suggests its potential as an effective strategy to bridge this gap and promote widespread Lp(a) testing and case identification, thereby enhancing care quality and patient outcomes on a larger scale.

In addition to the extremely low uptake of Lp(a) testing, our findings suggest that the subset of individuals currently undergoing Lp(a) testing is distinctively selective. This population is quantitatively different from population-based cohorts and even a randomly selected sample of patients from the EHR of the same health system. This could stem from healthcare providers preferentially ordering Lp(a) tests for high-risk patients or those with recurrent cardiovascular events.^15–17^ Moreover, the gradual dissemination of new medical practices and the potential future availability of targeted therapeutics may catalyze the broader adoption of universal Lp(a) screening. However, there is a wide gap between existing knowledge and its practical application, wherein ARISE implementation within health systems represents a promising avenue to address this disconnect.

Our findings support leveraging ARISE for large-scale Lp(a) screening within health systems, which will lead to increased Lp(a) testing and identification of those with elevated Lp(a). These individuals should subsequently undergo rigorous cardiovascular risk management, e.g., statins, proprotein convertase subtilisin/kexin type 9 inhibitors, and lifestyle modifications.^6,15–17,21–25^ Additionally, novel Lp(a)-targeted medications are being developed and tested in randomized clinical trials.^9–14^ Nevertheless, inadequate testing challenges the timely recruitment of the participants. Therefore, case finding through health system screening can accelerate trial enrollment and discoveries.

The study findings should be interpreted in the context of its limitations. First, this is a retrospective study assessing the feasibility of ARISE deployment within health systems. Therefore, future prospective studies where Lp(a) testing is informed by ARISE risk stratification should be conducted as the next step. Second, we defined study covariates, including past medical and medication history, relative to the Lp(a) or LDL-C measurement data using EHR. While this may introduce bias due to the real-world nature of EHR compared with a strictly controlled research environment, we employed tailored definitions to ensure complete capture of relevant information. Furthermore, the measurement date of LDL-C, the most important ARISE feature, was chosen as the index date for computing the ARISE score in individuals not tested for Lp(a), to enhance the robustness of this screening strategy. Third, the recruitment period of US cohorts was a decade earlier than that of other study cohorts when indications of statin use were not as broad. This could affect Gower’s distance between the US cohorts and the YNHHS Lp(a) cohort. However, the observed differences between the UKB and a randomly selected YNHHS sample, when compared with the YNHHS Lp(a) cohorts, underscore the systematic variance in populations undergoing Lp(a) testing. Fourth, all screening strategies, including ARISE deployment in health systems, are associated with false positives and negatives. Nonetheless, there is an escalating endorsement for universal Lp(a) assessments across the entire population.^6^ Hence, ARISE can serve as an easily deployable, algorithmic strategy to achieve this goal, despite the presence of false positives and negatives. Additionally, we have previously demonstrated that undergoing Lp(a) testing does not affect adverse cardiovascular outcomes for individuals with a negative ARISE score, underscoring that the implementation of ARISE does not lead to worse outcomes in those not advised to undergo testing.^18^

## CONCLUSIONS

We demonstrate that the deployment of ARISE, a novel algorithmic strategy to define the risk of elevated Lp(a), is feasible based on already available data for patients in US health systems, enabling a high-yield testing strategy. We also confirm the very low adoption of Lp(a) testing conducted in a highly selective population, underscoring the need for an algorithmically enriched screening strategy.

## Supporting information

Supplemental Content

## Data Availability

The analyzed de-identified data are available for the UK Biobank cohort from the UK Biobank's Access Management System and for ARIC, CARDIA, and MESA cohorts from the NHLBI's Biologic Specimen and Data Repository Information Coordinating Center (BioLINCC). The analyzed data from Yale-New Haven Health System and Vanderbilt University Medical Center cannot be made publicly available.

## ACKNOWLEDGEMENTS

The American Heart Association Precision Medicine Platform (https://precision.heart.org/) was used for data analysis. The dataset from Vanderbilt University Medical Center’s BioVU is supported by numerous sources: institutional funding, private agencies, and federal grants. These include the NIH funded Shared Instrumentation Grant S10RR025141; and CTSA grants UL1TR002243, UL1TR000445, and UL1RR024975.

## Non-standard Abbreviations and Acronyms

ARIC: Atherosclerosis Risk in Communities

ARISE: Algorithmic Risk Inspection for Screening Elevated Lp(a)

ASCVD: atherosclerotic cardiovascular disease

CARDIA: Coronary Artery Risk Development in Young Adults

EHR: electronic health records

HDL-C: high-density lipoprotein cholesterol

LDL-C: low-density lipoprotein cholesterol

Lp(a): Lipoprotein(a)

MESA: Multi-Ethnic Study of Atherosclerosis

UKB: UK Biobank

UMAP: uniform manifold approximation and projection

VUMC: Vanderbilt University Medical Center

YNHHS: Yale-New Haven Health System

