## Supplemental Content for "Evaluation of a Machine Learning-guided Strategy for Elevated Lipoprotein(a) Screening in Health Systems"

### **SUPPLEMENTAL METHODS**

**Data Sources**

We included data from two US health systems, the Yale-New Haven Health System (YNHHS) and the Vanderbilt University Medical Center (VUMC). To compare the representation of the Lipoprotein(a) [Lp(a)]-tested populations in these health systems with population-based cohorts, we included one cohort from the UK, the UK Biobank (UKB), and three cohorts from the US, Atherosclerosis Risk in Communities (ARIC), Coronary Artery Risk Development in Young Adults (CARDIA) and Multi-Ethnic Study of Atherosclerosis (MESA).

YNHHS provides care to more than 2.7 million patients through five acute-care hospitals (Yale New Haven Hospital, Bridgeport Hospital, Greenwich Hospital, Lawrence + Memorial Hospital, and Westerly Hospital), and a multispecialty network of outpatient clinics (Northeast Medical Group) across Connecticut, New York, and Rhode Island. We included two populations from the YNHHS during 2013-2023: (1) 7,981 patients who underwent at least one Lp(a) assessment with a valid serum Lp(a) value in the YNHHS electronic health records (EHR), and (2) a random sample of 100,000 patients who had at least one encounter in the YNHHS EHR.

VUMC provides care to over 3.6 million patients through VUMC hospitals and a network of owned primary-care clinics across Tennessee and Kentucky. We included 10,635 patients with at least one Lp(a) assessment with a valid serum Lp(a) value in the VUMC EHR from 1997-2022. The dataset was de-identified and mapped to the Observational Health Data Sciences & Informatics’ Observational Medical Outcomes Partnership (OMOP) common data model by Nashville Biosciences.

Among the prospective cohorts, UKB recruited people aged 40-69 years in the UK during 2006-2010. We included 456,815 UKB participants who had undergone Lp(a) assessment on their first visit in 2006-2010 or their second visit in 2012-2013.^1^ The ARIC study included 14,484 participants aged 45-64 years from four US communities whose Lp(a) were tested in 1987-1989.^2^ The CARDIA study included 4,124 participants aged 18-30 years recruited from four US urban areas in 1985-1986 who had undergone Lp(a) assessment in 1990-1991.^3^ From the MESA study, we included 4,672 participants aged 45-84 years from six centers whose Lp(a) were tested in 2000-2002.^4^ We have presented the results of ARIC, CARDIA, and MESA cohorts pooled together as a single US cohort.

**Study Outcome**

We assessed the feasibility of deploying Algorithmic Risk Inspection for Screening Elevated Lp(a) (ARISE) by calculating the proportion of patients in a random sample from YNHHS who had not undergone Lp(a) testing but for whom the ARISE score, and therefore, a probability of elevated Lp(a) could be computed using data already available in the EHR. Furthermore, we evaluated the association of a positive ARISE score (ARISE score ≥0.203 as defined in the development cohort) with elevated Lp(a) (≥150 nmol/L) among the smaller subset that had undergone Lp(a)-testing at both YNHHS and VUMC.^5^

In addition, we quantified the differences between populations tested for Lp(a) and other study groups using a multidimensional distance metric – Gower’s distance. Gower’s distance is computed as a pairwise distance metric between two individuals, ranging from 0 for an identical pair on all measured characteristics (both categorical and continuous) and 1 for the most dissimilar, with a majority of values ranging between 0 and 1.^6^ We calculated the Gower’s dissimilarity distances for each pair of the study cohorts based on the six ARISE features: (1) atherosclerotic cardiovascular disease (ASCVD) history, (2) current use of statins, (3) current use of anti-hypertensive medications, (4) serum low-density lipoprotein cholesterol (LDL-C), (5) serum high-density lipoprotein cholesterol (HDL-C), and (6) serum triglycerides.^5^ To quantify the heterogeneity between each pair of the study cohorts, we calculated the median of the medians of Gower's distances for each pair. We first calculated the median of Gower's distances between each individual in one cohort relative to all individuals in the other cohort. To summarize the overall dissimilarity between the cohorts, we then reported the median of these individual medians. This measure, the median of the medians of Gower's distances, offers a robust metric of the central dissimilarity between the two groups.^7^

**Study Covariates**

We extracted several key elements across all cohorts, including demographics (age, sex, and race and ethnicity) to characterize the study population and the six input features of ARISE for comparing the study cohorts. For patients in the health system, the index date for evaluating ARISE features was defined as (1) the date of their first Lp(a) assessment in the Lp(a) tested population, (2) their first LDL-C assessment date if the patient represented the study group not tested for Lp(a). The choice of LDL-C assessment date as the index date, as opposed to other ARISE features, stems from the fact that LDL-C was the most important feature in ARISE in the development and external validation cohorts.^5^ ASCVD history was defined as the presence of relevant diagnosis or procedural codes in the EHR from any time before and up to 30 days after the index date. We identified ASCVD history using a combination of diagnosis codes defined by International Classification of Diseases, 9^th^ Revision, Clinical Modification (ICD-9-CM), and ICD-10-CM, and procedure codes defined by International Classification of Diseases, 9^th^ Revision, Procedure Coding System (ICD-9-PCS), ICD-10-PCS, and Current Procedural Terminology (CPT-4).

The diagnosis codes included ischemic heart disease, ischemic stroke, and peripheral arterial disease, and the procedural codes spanned all coronary, cerebral, and peripheral arterial revascularization procedures (**Table S1**). The concurrent use of medications – specifically statins and anti-hypertensive medications - was defined as the presence of any prescription records for the medication from one year before and up to the index date (list of medications in **Table S2**). Lab values obtained from 30 days before and up to the index date were included, with the date defined by the testing date as opposed to the reporting date.

For the population-based cohorts of UKB, ARIC, CARDIA, and MESA, the study covariates have been defined based on the Lp(a) assessment date in each cohort.^5^ The study covariates comprised demographics (age, sex, and race and ethnicity) and the six input features of ARISE, including ASCVD history, use of statins, use of anti-hypertensive medications, serum LDL-C, serum HDL-C, and serum triglycerides.

UKB has a unique universal linkage of EHR for all of its participants encompassing historical data prior to enrollment in the UKB. This included data from the Hospital Episode Statistics for England starting from 1997, the Scottish Morbidity Record beginning in 1981, and the Patient Episode Database for Wales from 1998 onwards. In the linked EHR, diagnosis codes were recorded using International Classification of Diseases, 9^th^ Revision (ICD-9) and ICD-10, and procedure codes were coded using Office of Population Censuses and Surveys Classification of Interventions and Procedures, versions 3 (OPCS-3) and OPCS-4. ASCVD history was defined using a combination of diagnosis and procedure codes (**Table S3**). We identified the use of statins and anti-hypertensive medications using the medication list self-reported by the participants (**Table S4**). Serum LDL-C, HDL-C, and triglycerides measured concurrently with Lp(a) were included.

In the US-based cohorts of ARIC, CARDIA, and MESA, ASCVD history was defined as a history of ischemic heart disease, ischemic stroke, peripheral arterial disease, coronary artery bypass grafting, percutaneous coronary intervention, carotid revascularization, or peripheral arterial revascularization self-reported by the participants. The use of statins and anti-hypertensive medications was self-reported by the participants. We included measurements of serum LDL-C, HDL-C, and triglycerides conducted in the same study visit as Lp(a).

**Statistical Analysis**

Continuous and categorical variables were reported as median [interquartile range (IQR)], and number and percentages, respectively. ARISE includes a data processing pipeline, which imputes missing data and scales continuous variables, and an extreme gradient boosting (XGBoost) model using the six features mentioned above. ARISE score, the probability of elevated Lp(a), was computed by deploying the data processing pipeline followed by the XGBoost model. We calculated the ARISE score for patients who had already available EHR data for all ARISE features. The data processing pipeline of ARISE has been developed and validated in large, multinational, prospective, population-based cohorts, representing a robust algorithm for imputing missing data.^5^ Therefore, as a sensitivity analysis, we also leveraged this pipeline to calculate the ARISE score for a subset of patients missing one or two data points for ARISE features.

In the populations defined within the YNHHS and VUMC who underwent Lp(a) testing, the association of ARISE score with elevated Lp(a) was assessed by fitting a logistic regression model with elevated Lp(a) as the binary dependent variable and the ARISE score as the binary independent variable. The threshold for ARISE was set at 0.203 for optimizing specificity at 90%, as described previously.^5^ We also reported the odds ratio (OR) of elevated Lp(a) per one standard deviation (SD) increase of the ARISE score. Additionally, we assessed the differences between each pairwise comparison of the study cohorts using Gower's dissimilarity distance, as described above.^6^ We also employed a dimensionality reduction technique, uniform manifold approximation and projection (UMAP), to visualize the relationships between the cohort participants.^8^ Statistical analyses were conducted using Python 3.11.2 and R version 4.2.0, employing two-sided statistical tests with the significance level set at 0.05. A secure workspace on the American Heart Association Precision Medicine Platform was used for data analysis.

### **Table S1. Definition of Atherosclerotic Cardiovascular Disease Using Diagnosis and Procedure Codes in the US Health Systems**

| **Condition/Procedure** | **Dictionary** | | |
| --- | --- | --- | --- |
| **Condition** | **ICD-9-CM Code** | **ICD-10-CM Code** | |
| Ischemic Heart Disease | 4100, 41000, 41001, 41002, 4101, 41010, 41011, 41012, 4102, 41020, 41021, 41022, 4103, 41030, 41031, 41032, 4104, 41040, 41041, 41042, 4105, 41050, 41051, 41052, 4106, 41060, 41061, 41062, 4107, 41070, 41071, 41072, 4108, 41080, 41081, 41082, 4109, 41090, 41091, 41092, 4110, 4111, 4118, 41181, 41189, 412, 4130, 4131, 4139, 4140, 41400, 41401, 41406, 4142, 4143, 4144, 4148, 4149, V4581, V4582 | I200, I201, I202, I208, I209, I237, I25110, I25111, I25112, I25118, I25119, I25700, I25701, I25702, I25708, I25709, I25710, I25711, I25712, I25718, I25719, I25720, I25721, I25722, I25728, I25729, I25730, I25731, I25732, I25738, I25739, I25750, I25751, I25752, I25758, I25759, I25760, I25761, I25762, I25768, I25769, I25790, I25791, I25792, I25798, I25799, I25810, I2101, I2102, I2109, I2111, I2119, I2121, I2129, I213, I214, I219, I21A1, I21A9, I220, I221, I222, I228, I229, I230, I231, I232, I233, I234, I235, I236, I237, I238, I240, I241, I248, I249, I2510,I25110, I25111, I25112, I25118, I25119, I252, I253, I2541, I2542, I255, I256, I25700, I25701, I25702, I25708, I25709, I25710, I25711, I25712, I25718, I25719, I25720, I25721, I25722, I25728, I25729, I25730, I25731, I25732, I25738, I25739, I25750, I25751, I25752, I25758, I25759, I25760, I25761, I25762, I25768, I25769, I25790, I25791, I25792, I25798, I25799, I25810, I25811, I25812, I2582, I2583, I2584, I2589, I259 | |
| Ischemic Stroke | 34660, 34661, 34662, 34663, 430, 431, 4320, 4321, 4329, 43301, 43311, 43321, 43331, 43381, 43391, 4340, 43400, 43401, 4341, 43410, 43411, 4349, 43490, 43491, 436, 4330, 43300, 4331, 43310, 4332, 43320, 4333, 43330, 4338, 43380, 4339, 43390 | G43601, G43609, G43611, G43619, I6300, I63011, I63012, I63013, I63019, I6302, I63031, I63032, I63033, I63039, I6309, I6310, I63111, I63112, I63113, I63119, I6312, I63131, I63132, I63133, I63139, I6319, I6320, I63211, I63212, I63213, I63219, I6322, I63231, I63232, I63233, I63239, I6329, I6330, I63311, I63312, I63313, I63319, I63321, I63322, I63323, I63329, I63331, I63332, I63333, I63339, I63341, I63342, I63343, I63349, I6339, I6340, I63411, I63412, I63413, I63419, I63421, I63422, I63423, I63429, I63431, I63432, I63433, I63439, I63441, I63442, I63443, I63449, I6349, I6350, I63511, I63512, I63513, I63519, I63521, I63522, I63523, I63529, I63531, I63532, I63533, I63539, I63541, I63542, I63543, I63549, I6359, I636, I638, I6381, I6389, I639, I6501, I6502, I6503, I6509, I651, I6521, I6522, I6523, I6529, I658, I659, I6601, I6602, I6603, I6609, I6611, I6612, I6613, I6619, I6621, I6622, I6623, I6629, I663, I668, I669, I672, I6781, I6782, I6930, I6931, I69310, I69311, I69312, I69313, I69314, I69315, I69318, I69319, I69320, I69321, I69322, I69323, I69328, I69331, I69332, I69333, I69334, I69339, I69341, I69342, I69343, I69344, I69349, I69351, I69352, I69353, I69354, I69359, I69361, I69362, I69363, I69364, I69365, I69369, I69390, I69391, I69392, I69393, I69398, I6980, I6981, I69810, I69811, I69812, I69813, I69814, I69815, I69818, I69819, I69820, I69821, I69822, I69823, I69828, I69831, I69832, I69833, I69834, I69839, I69841, I69842, I69843, I69844, I69849, I69851, I69852, I69853, I69854, I69859, I69861, I69862, I69863, I69864, I69865, I69869, I69890, I69891, I69892, I69893, I69898, I6990, I6991, I69910, I69911, I69912, I69913, I69914, I69915, I69918, I69919, I69920, I69921, I69922, I69923, I69928, I69931, I69932, I69933, I69934, I69939, I69941, I69942, I69943, I69944, I69949, I69951, I69952, I69953, I69954, I69959, I69961, I69962, I69963, I69964, I69965, I69969, I69990, I69991, I69992, I69993, I69998 | |
| Peripheral Arterial Disease | 4400, 4401, 4402, 44020, 44021, 44022, 44023, 44029, 4404, 4408, 4409, 4439, 5570,  5571, 5579 | I70201, I70202, I70203, I70208, I70209, I70211, I70212, I70213, I70218, I70219, I70221, I70222, I70223, I70228, I70229, I70231, I70232, I70233, I70234, I70235, I70238, I70239, I70241, I70242, I70243, I70244, I70245, I70248, I70249, I7025, I70261, I70262, I70263, I70268, I70269, I70291, I70292, I70293, I70298, I70299, I70301, I70302, I70303, I70308, I70309, I70311, I70312, I70313, I70318, I70319, I70321, I70322, I70323, I70328, I70329, I70331, I70332, I70333, I70334, I70335, I70338, I70339, I70341, I70342, I70343, I70344, I70345, I70348, I70349, I7035, I70361, I70362, I70363, I70368, I70369, I70391, I70392, I70393, I70398, I70399, I70401, I70402, I70403, I70408, I70409, I70411, I70412, I70413, I70418, I70419, I70421, I70422, I70423, I70428, I70429, I70431, I70432, I70433, I70434, I70435, I70438, I70439, I70441, I70442, I70443, I70444, I70445, I70448, I70449, I7045, I70461, I70462, I70463, I70468, I70469, I70491, I70492, I70493, I70498, I70499, I70501, I70502, I70503, I70508, I70509, I70511, I70512, I70513, I70518, I70519, I70521, I70522, I70523, I70528, I70529, I70531, I70532, I70533, I70534, I70535, I70538, I70539, I70541, I70542, I70543, I70544, I70545, I70548, I70549, I7055, I70561, I70562, I70563, I70568, I70569, I70591, I70592, I70593, I70598, I70599, I70601, I70602, I70603, I70608, I70609, I70611, I70612, I70613, I70618, I70619, I70621, I70622, I70623, I70628, I70629, I70631, I70632, I70633, I70634, I70635, I70638, I70639, I70641, I70642, I70643, I70644, I70645, I70648, I70649, I7065, I70661, I70662, I70663, I70668, I70669, I70691, I70692, I70693, I70698, I70699, I70701, I70702, I70703, I70708, I70709, I70711, I70712, I70713, I70718, I70719, I70721, I70722, I70723, I70728, I70729, I70731, I70732, I70733, I70734, I70735, I70738, I70739, I70741, I70742, I70743, I70744, I70745, I70748, I70749, I7075, I70761, I70762, I70763, I70768, I70769, I70791, I70792, I70793, I70798, I70799, I7092, I739, I742, I743, I744, I745, I748, I749, I75011, I75012, I75013, I75019, I75021, I75022, I75023, I75029, I7581, I7589 | |
| **Procedure** | **ICD-9-PCS** | **ICD-10-PCS** | **CPT-4** |
| Coronary Artery Bypass Grafting | 3610, 3611, 3612, 3613, 3614, 3615, 3616, 3617, 3619, 362, 363, 3631, 3632, 3633, 3634, 3639 | 02100, 02104, 02110, 02114, 02120, 02124, 02130, 02134 | 33510, 33511, 33512, 33513, 33514, 33516, 33533, 33534, 33535, 33536, 33517, 33518, 33519, 33521, 33522, 33523, 33530, 33572 |
| Percutaneous Coronary Intervention | 0066, 1755, 3601, 3602, 3605, 3604 | 02103, 02113, 02123, 02133, 02703, 02713, 02723, 02733, 02C03, 02C13, 02C23, 02C33, 02F03, 02F13, 02F23, 02F33, 02H03, 02H13, 02H23, 02H33, 02N03, 02N13, 02N23, 02N33, 02Q03, 02Q13, 02Q23, 02Q33, 02U03, 02U13, 02U23, 02U33, 5A0222C, X2C03, X2C13, X2C23, X2C33 | 92920, 92921, 92924, 92925, 92928, 92929, 92933, 92934, 92937, 92938, 92941, 92943, 92944, 92973, 92975, C9600, C9601, C9602, C9603, C9604, C9605, C9606, C9607, C9608 |
| Carotid Revascularization | 3811, 3812 | 031H0, 031J0, 031K0, 031L0, 031M0, 031N0, 037H0, 037H3, 037H4, 037J0, 037J3, 037J4, 037K0, 037K3, 037K4, 037L0, 037L3, 037L4, 037M0, 037M3, 037M4, 037N0, 037N3, 037N4, 03CH0, 03CH3, 03CH4, 03CJ0, 03CJ3, 03CJ4, 03CK0, 03CK3, 03CK4, 03CL0, 03CL3, 03CL4, 03CM0, 03CM3, 03CM4, 03CN0, 03CN3, 03CN4, 03QH0, 03QH3, 03QH4, 03QJ0, 03QJ3, 03QJ4, 03QK0, 03QK3, 03QK4, 03QL0, 03QL3, 03QL4, 03QM0, 03QM3, 03QM4, 03QN0, 03QN3, 03QN4, 03RH0, 03RH4, 03RJ0, 03RJ4, 03RK0, 03RK4, 03RL0, 03RL4, 03RM0, 03RM4, 03RN0, 03RN4, 03UH0, 03UH3, 03UH4, 03UJ0, 03UJ3, 03UJ4, 03UK0, 03UK3, 03UK4, 03UL0, 03UL3, 03UL4, 03UM0, 03UM3, 03UM4, 03UN0, 03UN3, 03UN4, 03VH0, 03VH3, 03VH4, 03VJ0, 03VJ3, 03VJ4, 03VK0, 03VK3, 03VK4, 03VL0, 03VL3, 03VL4, 03VM0, 03VM3, 03VM4, 03VN0, 03VN3, 03VN4, X2AH336, X2AJ336 | 37215, 37216, 37217, 37218 |
| Peripheral Arterial Revascularization | 3925, 3929, 3808, 3818 | 0410090, 04104, 041C0, 041C4, 041D0, 041D4, 041E0, 041E4, 041F0, 041F4, 041H0, 041H4, 041J0, 041J4, 041K0, 041J3, 041K4, 041L0, 041L3, 041L4, 041M0, 041M3, 041M4, 041N0 041N3, 041N4, 041P0, 041P3, 041P4, 041Q0, 041Q3, 041Q4, 041R0, 041R3, 041R4, 041S0, 041S3, 041S4, 041T0, 041T3, 041T4, 041U0, 041U3, 041U4, 041V0, 041V3, 041V4, 041W0, 041W3, 041W4 | 37220, 37221, 37222, 37223, 37224, 37225, 37226, 37227, 37228, 37229, 37230, 37231, 37232, 37233, 37234, 37235 |

**Abbreviations:** ICD-9-CM, International Classification of Diseases, 9th Revision, Clinical Modification; ICD-9-PCS, International Classification of Diseases, 9th Revision, Procedure Coding System; CPT-4, Current Procedural Terminology.

### **Table S2. Medication Lists for Statins and Anti-hypertensive Medications in the US Health Systems**

| **Medication Category** | **Medication Name** |
| --- | --- |
| Statin | Atorvastatin, Rosuvastatin, Pravastatin, Lovastatin, Simvastatin, Pitavastatin, Fluvastatin, Cerivastatin |
| Anti-hypertensive Medications | Valsartan, Losartan, Olmesartan, Candesartan, Irbesartan, Telmisartan, Eprosartan, Azilsartan,  Lisinopril, Ramipril, Benazepril, Quinapril, Enalapril, Fosinopril, Perindopril, Captopril, Trandolapril, Moexipril, Enalaprilat,  Nicardipine, Amlodipine, Felodipine, Nifedipine, Clevidipine, Nisoldipine, Isradipine,  Doxazosin, Prazosin, Terazosin, Alfuzosin,  Carvedilol, Metoprolol, Atenolol, Nadolol, Bisoprolol, Nebivolol, Pindolol, Acebutolol, Penbutolol,  Hydrochlorothiazide, Chlorothiazide, Cyclothiazide, Bendroflumethiazide, Trichlormethiazide, Chlorthalidone, Indapamide, Xipamide, Metolazone,  Amiloride, Triamterene, Spironolactone, Eplerenone |

### **Table S3. Definition of Atherosclerotic Cardiovascular Disease Using Diagnosis and Procedure Codes in the UK Biobank**

| **Condition/Procedure** | **Dictionary** | |
| --- | --- | --- |
| **Condition** | **ICD-9 Code** | **ICD-10 Code** |
| Ischemic Heart Disease | 410, 4109, 411, 4119, 412, 4129, 413, 4139, 414, 4140, 4148, 4149 | I20, I200, I208, I209, I21, I210, I211, I212, I213, I214, I219, I21X, I22, I220, I221, I228, I229, I23, I230, I231, I232, I233, I234, I235, I236, I238, I24, I240, I241, I248, I249, I25, I250, I251, I252, I255, I256, I258, I259, Z951, Z955 |
| Ischemic Stroke | 433, 4330, 4331, 4332, 4333, 4338, 4339, 434, 4340, 4341, 4349, 435, 4359, 437, 4370, 4371 | G45, G450, G451, G452, G453, G454, G458, G459, I63, I630, I631, I632, I633, I634, I635, I638, I639, I64, I65, I650, I651, I652, I653, I658, I659, I66, I660, I661, I662, I663, I664, I668, I669, I672, I693, I694 |
| Peripheral Arterial Disease | 4402, 4442 | I702, I7020, I7021, I742, I743, I744 |
| **Procedure** | **OPCS-3 Code** | **OPCS-4 Code** |
| Coronary Artery Bypass Grafting | 3043 | K40, K401, K402, K403, K404, K408, K409, K41, K411, K412, K413, K414, K418, K419, K42, K421, K422, K423, K424, K428, K429, K43, K431, K432, K433, K434, K438, K439, K44, K441, K442, K448, K449, K45, K451, K452, K453, K454, K455, K456, K458, K459, K46, K461, K462, K463, K464, K465, K468, K469 |
| Percutaneous Coronary Intervention | None | K49, K491, K492, K493, K494, K498, K499, K50, K501, K502, K503, K504, K508, K509, K75, K751, K752, K753, K754, K758, K759 |
| Carotid Revascularization | None | L29, L291, L292, L293, L294, L295, L296, L297, L298, L299, L303, L31, L311, L313, L314, L318, L319 |
| Peripheral Arterial Revascularization | 881, 8811 | L50, L501, L502, L503, L504, L505, L506, L508, L509, L51, L511, L512, L513, L514, L515, L516, L518, L519, L52, L521, L522, L528, L529, L532, L54, L541, L542, L544, L548, L549, L58, L581, L582, L583, L584, L585, L586, L587, L588, L589, L59, L591, L592, L593, L594, L595, L596, L597, L598, L599, L60, L601, L602, L603, L604, L608, L609, L622, L63, L631, L632, L633, L635, L638, L639, L66, L661, L662, L665, L667, L681, L682, L701, L71, L711, L712, L713, L714, L715, L716, L717, L718, L719 |

**Abbreviations:** ICD, International Classification of Diseases; OPCS, Office of Population Censuses and Surveys Classification of Interventions and Procedures.

### **Table S4. Medication Lists for Statins and Anti-hypertensive Medications in the UK Biobank**

| **Medication Category** | **Medication Name** |
| --- | --- |
| Statin | Simvastatin, Velastatin, Zocor, Simvador, Synvinolin, Fluvastatin, Lescol, Pravastatin, Eptastatin, Lipostat, Atorvastatin, Lipitor, Rosuvastatin, Crestor |
| Anti-hypertensive Medications | Provided by the UK Biobank as a Variable |
